# Risk factors, early presentations, and clinical markers of Parkinson’s disease in Primary Care in a diverse UK population

**DOI:** 10.1101/2021.09.30.21264336

**Authors:** Cristina Simonet, Jonathan Bestwick, Mark Jitlal, Aaron Ben-Joseph, Charles R. Marshall, Ruth Dobson, Soha Marrium, John Robson, Andrew J. Lees, Gavin Giovannoni, Jack Cuzick, Anette Schrag, Alastair J. Noyce

## Abstract

**Importance:** Predictors of future Parkinson’s disease (PD) have been suggested through population-based studies, but these studies over-represent white, affluent groups and may not be generalisable.

**Objective:** To investigate the association between risk factors and pre-diagnostic presentations of PD in a uniquely diverse UK population, with universal access to health care.

**Design, Setting, and Participants:** A case-control study was conducted in East London, using primary care health records.

**Main outcomes and Measures:** Logistic regression was used to determine associations between risk factors and pre-diagnostic presentations of PD with final diagnosis. Three periods (recorded <2 years, 2-5 years, and 5-10 years prior to diagnosis) were analysed.

**Results:** Primary care records were available for 1,055 patients with PD and 1,009,523 controls. The strongest associations were found for tremor (OR, 181.69; 95% CI, 151.91-217.31) and ‘memory complaints’ (OR, 9.84; 95% CI, 7.39-13.11), <2 years before PD diagnosis. However, associations for both complaints persisted up to 10 years prior to PD diagnosis. Shoulder pain was more common in those who developed PD, emerging 5 to 10 years prior to diagnosis (OR, 2.54; 95% CI, 1.77-3.65), and may be a surrogate marker for rigidity. Epilepsy showed a strongest association with subsequent PD (OR, 5.14; 95% CI, 1.26-21.0), and associations were also found for hypertension (OR, 1.71; 95% CI, 1.34-2.17) and type 2 diabetes (OR, 1.57; 95% CI, 1.31-1.87) 5 to 10 years before diagnosis. We replicated several known associations with early non motor features including hypotension, constipation, and depression. A weak but novel association was observed with prodromal hearing loss and subsequent PD, which appeared up to 10 years prior to diagnosis. No associations with future PD diagnosis were found for ethnicity or deprivation index.

**Conclusions and relevance:** This study provides further evidence that a range of comorbidities and pre-diagnostic presentations are encountered in primary care prior to PD diagnosis, but for the first time in such a diverse and deprived population. Convincing temporal associations were observed for epilepsy and hearing loss with subsequent PD. The predominance of ‘memory symptoms’ hints at an excess of cognitive dysfunction in early PD in this population or difficulty in correctly ascertaining symptoms in traditionally under-represented groups.

**Key points:** *Question:* What are the pre-diagnostic manifestations of Parkinson’s disease (PD) that present to primary care in a diverse and deprived population with universal access?

*Findings:* Tremor and ‘memory symptoms’ were reported up to 10 years before diagnosis and were most strongly associated with PD. Several recognised pre-diagnostic features were replicated, but novel temporal associations between epilepsy and hearing loss with subsequent PD were observed.

*Meaning:* The range of symptoms that cause people to present to primary care, up to a decade before PD diagnosis, are broad. The strength of association, relative importance and interpretation of prodromal features may vary according to the population studied.

## Introduction

Diversity is lacking in the study of complex diseases, including Parkinson’s disease (PD) and research has been conducted in patients of Northern European ancestry *[1,2]*. As such, little is known about how PD manifests in different ethnic groups, and whether there is differential case ascertainment, response to medication, or different determinants of risk and phenotype *[3,4]*. It is not only patients from minority ethnic groups who are under-represented in PD research *[5]*. Very little is known about PD occurring in people living in areas of high deprivation, with high unemployment, social isolation, and those residing in impoverished urban settings *[6]*. Geographical location might be an important determinant of PD risk *[7,8]*. The National Health Service provides primary and secondary care to the entire UK population, free at the point of access. Publicly funded healthcare systems such as the NHS are ideal for the study of disease determinants that may vary according to the above factors, to reduce bias and improve generalisability of results.

Early features, risk factors and predictors of PD diagnosis have been suggested through large, population-based, observational studies *[10–14]*. In the UK, a comprehensive observational study of early features of PD was undertaken by some members of this study group in the THIN primary care database that involved 8,166 people with PD and 46,455 healthy controls, who were mainly White and came from higher-income groups*[12]*. In the present study, we used a similar approach, but in a highly diverse population from East London, with some of the highest levels of deprivation in the UK.

## Methods

### Study design

This is a case-control study, nested in a large primary care dataset in East London. Primary care data was compiled from searches of EMIS (Egton Medical Information Systems) electronic healthcare records system for the SHARE project (Secure Health Analysis and Research in East London). The database included health records of 1,016,277 patients from General Practices across four Clinical Commissioning Groups (CCGs) in East London: Hackney & City of London, Newham, Tower Hamlets, and Waltham Forest. EMIS was begun in the UK in 1990 and paper records acquired prior to this were manually transcribed into the system.

In the UK National Health System (NHS), each individual is identified by a unique NHS number which enables linkage to their medical records. When individuals move between healthcare providers their records move with them. All non-emergency secondary care referrals originate from primary care, and outcomes are communicated back, thus primary care records represent aggregated medical information about an individual throughout their life.

### Identification of cases and controls

All individuals with a code of “Parkinson’s disease” as a diagnosis were included as cases in the analysis. Patients with PD but missing a date of diagnosis were excluded as well as those with a diagnosis of dementia, atypical PD, and other neurodegenerative conditions. Controls were those without a code of “Parkinson’s disease” or other chronic neurological conditions such as dementia, multiple sclerosis, atypical Parkinson’s, and motor neurone disease. Epilepsy was not excluded since there was a prior interest to explore an association given two recent publications from members of our group in different settings (UK Biobank and 23andMe)*[13,14]*. Controls were assigned a “dummy date of PD diagnosis”, which was calculated as follows: the median age of PD diagnosis (69.0 years) was added to the year of birth of each control to create a dummy date of ‘diagnosis’ in controls. Then the earliest date between the “dummy date of PD diagnosis” and 6th February 2018, which was when the database was locked, was used as the time point to categorise the selected exposures as pre-diagnostic risk factors.

### Exposure selection and extraction

Exposures were selected based on a comprehensive meta-analysis on pre-diagnostic features and risk factors for PD, carried out by members of this group in 2012 *[15]*, and three other large studies of the pre-diagnostic phase of PD *[12,14,16]*. Together with epilepsy, hearing loss was also added since evidence had been recently published about its potential role as an early manifestation of PD *[17]*.

Overall, 27 variables were selected and subdivided into three categories: 1) comorbidities and risk factors, 2) pre-diagnostic non-motor manifestations (metabolic, sensory, autonomic, and neuropsychiatric), and 3) pre-diagnostic motor manifestations. Individual patient information was extracted by the Clinical Effectiveness Group (CEG) at Queen Mary University of London on 6th February 2018. All risk factors were recorded up to twice in the ELGP database (earliest ever record and the most recent record). Where there were repeat observations, the earliest date was used for the analysis.

Given the cross-sectional nature of data extraction, incidence rates could not be calculated. However, we wished to examine temporal relationships and so three periods of time were established to extract selected exposures (<2 years, 2-5 years, and 5-10 years before PD diagnosis/dummy diagnosis). We selected the same periods used in the THIN database study in order to make findings comparable and see whether there were differences between the two different populations with divergent socio-economic background and ethnicity *[12]*. Variables with less than 1% prevalence among PD cases across all 10-year periods were excluded from the analysis.

### Definition of Exposures

The variables extracted were based on identified data comprising diagnoses, laboratory results, and demographic details coded using the Read coding system (https://digital.nhs.uk/services/terminology-and-classifications/read-codes). Variables were defined so that as much data as possible could be used in the modelling. For this reason, missing data was categorised as “unknown” in the models rather than excluded.

#### Age

Age was taken as the age at data extraction (6th February 2018).

#### Ethnicity

Ethnicity was defined by the self-reported UK census categories, grouped here into major ethnic groups in the East London population: White (British, Irish, Other White), Black (African, Caribbean, Other Black), South Asian (Bangladeshi, Indian, Pakistani), other (Chinese, other and mixed groups) and unknown.

#### Index of Multiple Deprivation (IMD)

IMD is a global measure based on socio-economic terms, including income, employment, education, health, crime, housing, and environment. Raw IMD scores were assigned to deciles derived from national data and converted into quintiles. Quintile 1 (IMD 1-2) represented the most deprived 20% and quintile 5 (IMD 9-10) the least deprived 20%. IMD group 1-2 was used as the reference category in the analyses.

#### Vascular risk factors

Coded diagnoses of hypertension, cholesterol, and type 2 diabetes (T2D) were primarily defined on four levels depending on whether the risk factor was never recorded, first recorded prior to PD diagnosis (or dummy date of diagnosis for controls), first recorded after PD diagnosis, or unknown where the data were missing. For each risk factor, the status was determined by the earliest record, unless this was missing, in which case the status at the latest date was used. Total cholesterol levels were taken from the clinical data and were considered valid if they ranged from (0.5-50.0) mmol/L. Values >5.0 mmol/L were indicative of hypercholesterolaemia. Hypertension and T2D were recorded according to the presence of a coded morbidity record. Patients with a diagnosis of type 1 diabetes were recorded as “normal”. Although overweight is also considered a vascular risk factor, it is mentioned separately below.

#### Smoking

Smoking status was defined as being coded as a current, ex-smoker or never smoker, but time before PD diagnosis was not considered, given that smoking initiation in later life is rare. For that reason, data related to smoking will cover all pre-diagnostic periods.

#### Body Mass Index (BMI)

BMI was calculated from clinical data using height and weight measurements and categorised as follows: normal (BMI 20.0-24.9 kg/m2), underweight (10.0-19.9 kg/m2), and overweight (25.0-50.0 kg/m2). For height and weight, the ranges were 100-250 cm and 30-250 kg respectively. Where height or the calculated BMI was outside ranges, these data were deemed unlikely and were reclassified as unknown. In the same way, participants with missing BMI data were classified as unknown, unless they had a diagnosis of obesity recorded. In which case they were classified as overweight. Unlike overweight, underweight is not linked to coexistent vascular disease. We therefore listed it under a ‘metabolic prodrome’ category.

#### Other comorbidities

Hearing loss and epilepsy were defined by a coded diagnosis. In the case of hearing loss, a referral for assessment due to reported hearing difficulty was also included.

#### Non-motor or motor pre-diagnostic manifestations

Clinical symptoms of PD included coded non-motor features such as memory problems, depression, anxiety, fatigue, erectile dysfunction, shoulder pain, neck pain, and constipation. Motor features included rigidity, tremor, and balance difficulties. All were defined by recorded diagnosis or coded symptoms.

### Statistical modelling

For the periods from <2 years, 2– 5 years, and 5–10 years before the index date, the overall occurrence of pre-diagnostic symptoms was calculated as the absolute number and percentage. Logistic regression was used to estimate the odds ratio (OR) for PD and 95% confidence interval (CI) for each factor of interest. In the main analysis, the control group was not matched for age and sex to avoid introducing bias related to the matching process. A multivariable logistic regression model was created in which estimates for each exposure of interest were adjusted for age and sex. Due to ethnic and IMD variability in the East London population, a separate multivariable logistic analysis was performed accounting for ethnicity and IMD alone and in combination with age and sex. A separate matched case-control analysis was run as a sensitivity analysis by matching 10 controls for each case according to age (calendar year) and sex. We stratified the degree of association and, where significant, arbitrarily divided these into three categories: strong association (OR ≥ 5 or ≤0.2), moderate association (OR 2-5 or 0.2-0.5) and weak association (OR ≤ 2 or ≥0.5). All analyses were performed using R (v4.0.2) and Stata v15 (StatCorp, College Station, Texas).

## Results

### Demographic information

This nested case-control study included 1,055 patients with PD and 1,009,523 controls. Demographic information for cases and controls is summarised in Table 1. Patients with PD were more likely to be older (mean age for cases 72.9 years; controls 40.3 years) and male (cases 59.9% male; controls 51.2% male). No differences between patients and controls were found in Indices of Multiple Deprivation (IMD), but most participants (>75%) resided in the most deprived IMD quintile. The ethnicity of participants reflected the diversity of the local population in East London and was similar among PD cases and controls (cases: 51% White, 16% Black, 20% South Asian, 8% other; controls: 44% White, 13% Black, 21% South Asian, 11% other). Neither IMD or ethnicity were associated with a diagnosis of PD.

**Table 1.** Demographic information of PD cases and unmatched controls in East London primary care data.

|  |  |  |  |
| --- | --- | --- | --- |
|  | PD<br>N=1055 | Controls<br>N=1,009,523 | Prevalence (% PD<br>cases per group) |
| Age (years): Mean (SD) | 72.9 (11.3) | 40.3 (15.2) | NA |
| Sex: |  |  |  |
| Female, n (%) | 423 (40.1) | 492,647 (48.8) | 0.09 |
| Male, n (%) | 632 (59.9) | 516,862(51.2) | 0.12 |
| Ethnicity: |  |  |  |
| White | 537 (50.9) | 441,522 (43.7) | 0.12 |
| Black | 166 (15.7) | 134,629 (13.3) | 0.12 |
| South Asian | 208 (19.7) | 216,763 (21.5) | 0.10 |
| Other | 88 (8.3) | 114,503 (11.3) | 0.08 |
| Unknown | 56 (5.3) | 102,106 (10.1) | 0.05 |
| IMD: |  |  |  |
| 1-2 (most deprived) | 472 (44.7) | 453,747 (44.9) | 0.10 |
| 3-4 | 419 (39.7) | 424,944 (42.1) | 0.10 |
| 5-6 | 68 (6.4) | 76,773 (7.6) | 0.09 |
| 7-8 | 25 (2.4) | 19,258 (1.9) | 0.13 |
| 9-10(least deprived) | 16 (1.5) | 6,895 (0.7) | 0.23 |
| Unknown | 55 (5.2) | 27,906 (2.8) | 0.002 |

### Association of mild life risk factors and comorbidities

Table 2 summarises associations between comorbidities or risk factors and PD over three time periods. An adjusted model for age and sex showed that the strongest association was for epilepsy, across each of the three time periods analysed (<2 years: OR, 5.14; 95% CI, 1.26 to 20.99; 2-5 years: OR, 3.92; 95% CI, 1.24 to 12.39; 5-10 years: OR, 4.52; 95% CI, 1.99 to 10.24). For midlife vascular risk factors, having hypertension or T2D 5-10 years before diagnosis, but not closer to diagnosis, were associated with a higher odds of subsequent PD (hypertension: OR, 1.57; 95% CI, 1.31 to 1.87; T2D: OR, 1.71; 95% CI, 1.34 to 2.17). In contrast with being overweight, for which no association was seen, being underweight in the period closest to PD diagnosis (<2 years) was associated with a higher odds of PD (OR, 2.14; 95% CI, 1.01 to 4.54). There was no evidence for an association between high cholesterol and subsequent PD (OR, 1.00; 95% CI, 0.84 to 1.2). Smoking and alcohol consumption showed an inverse association when recorded further from PD diagnosis (5-10 years: smoking OR, 0.67; 95% CI, 0.5 to 0.88; alcohol OR, 0.75; 95% CI, 0.57 to 0.98) but no association when recorded closer to diagnosis (<2 years).

**Table 2.** Prevalence and adjusted odds ratios of PD for comorbidities and risk factors according to time of presentation.

| Exposures | Category | <2 years |  | 2-<5 years |  | 5-<10 years |  |
| --- | --- | --- | --- | --- | --- | --- | --- |
|  |  | %<br>(PD: Controls) | Adjusted OR<br>(95%CI) | %<br>(PD: Controls) | Adjusted OR<br>(95%CI) | %<br>(PD: Controls) | Adjusted OR<br>(95% CI) |
| **Overweight** | Lifestyle | 57 (5.4%):<br>65,775 (6.5%) | 1.12 (0.86 to<br>1.47) | 70 (6.6%): 83,536<br>(8.3%) | 0.82 (0.64 to<br>1.05) | 146 (13.8%): 106,421<br>(10.5%) | 1.15 (0.96 to<br>1.37) |
| **Smoking (ever)\*** |  | 361 (34.2%):<br>33,9129 (33.6%) | 0.82 (0.72 to<br>0.93) | 361 (34.2%): 33,9129<br>(33.6%) | 0.82 (0.72 to<br>0.93) | 361 (34.2%): 33,9129<br>(33.6%) | 0.82 (0.72 to<br>0.93) |
| **Alcohol (ever)** |  | 32 (3%): 67,994<br>(6.7%) | 0.86 (0.60 to<br>1.23) | 39 (3.7%): 89,842<br>(8.9%) | 0.73 (0.53 to<br>1.01) | 58 (5.5%): 88,696<br>(8.8%) | 0.75 (0.57 to<br>0.98) |
| **Type 2 diabetes** | Vascular<br>risk<br>factors | 27 (2.6%): 9,964<br>(1.0%) | 1.15 (0.78 to<br>1.69) | 34 (3.2%): 13,685<br>(1.4%) | 1.0 (0.71 to<br>1.41) | 74 (7.0%): 16,627<br>(1.6%) | 1.71 (1.34 to<br>2.17) |
| **Hypertension** |  | 48 (4.5%):<br>15,020 (1.5%) | 1.02 (0.76 to<br>1.37) | 69 (6.5%): 21,234<br>(2.1%) | 0.99 (0.77 to<br>1.26) | 143 (13.6%): 28,972<br>(2.9%) | 1.57 (1.31 to<br>1.87) |
| **High cholesterol** |  | 65 (6.2%):<br>39,087 (3.9%) | 1.13 (0.88 to<br>1.46) | 82 (7.8%): 57,876<br>(5.7%) | 0.87 (0.7 to 1.1) | 143 (13.6%): 80,205<br>(7.9%) | 1.0 (0.84 to 1.2) |
| **Epilepsy** | Other | 2 (0.2%): 603<br>(0.1%) | 5.14 (1.26 to<br>20.99) | 3 (0.3%): 875 (0.1%) | 3.92 (1.24 to<br>12.39) | 6 (0.6%):1,310 (0.1%) | 4.52 (1.99 to<br>10.24) |
| **Head injury** |  | 9 (0.9%): 4,120<br>(0.4%) | 3.21 (1.65 to<br>6.24) | 2 (0.2%): 4,693<br>(0.5%) | 0.62 (0.15 to<br>2.49) | 7 (0.7%): 5,718<br>(0.6%) | 1.82 (0.86 to<br>3.85) |

Standard: multivariable logistic model for PD with OR and 95% CI adjusted for age and sex. Time span: <2 years, 2-<5 years and 5-<10 years before PD diagnosis or index date. OR: Odds Ratio. CI: Confidence Interval. PD: Parkinson's disease patients (n=1055), controls (n=1,009,523). \*Data covers all pre-diagnostic period. It was not possible to classify data into 3 periods given that smoking initiation in later life is rare, therefore time before PD diagnosis was not considered.

Although ethnicity and IMD were not associated with PD, we adjusted for both covariates in addition to age and sex (Table S1 in the supplementary material), but there was no suggestion of confounding. The distribution of patients presenting with motor symptoms (tremor, rigidity, and balance difficulties) and ‘memory complaints’ in patients diagnosed with PD did not remarkably differ between ethnic groups (Table S2 in supplementary material).

### Pre-diagnostic manifestations

Table 3 summarises clinical manifestations in the three periods of time.

**Table 3.**
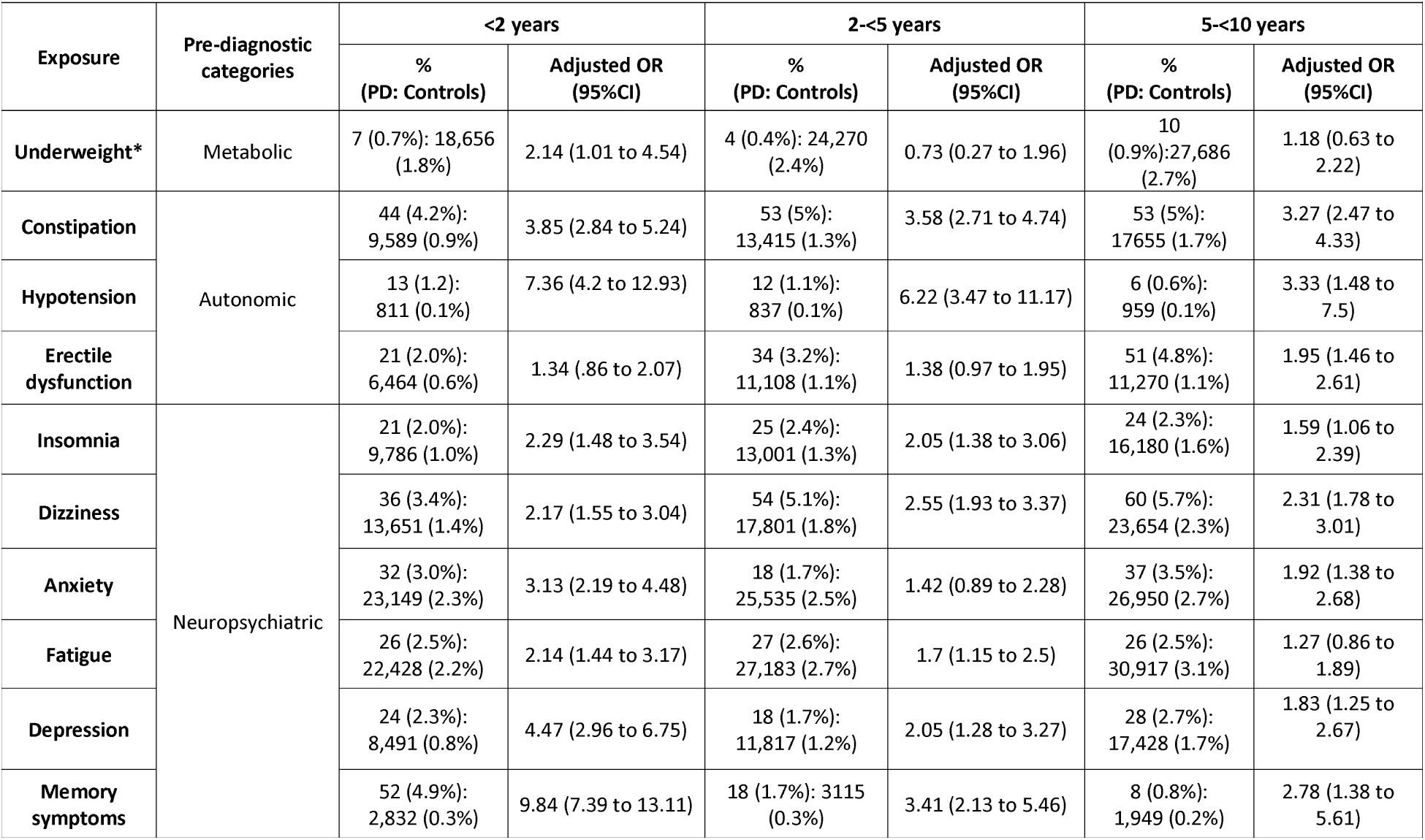
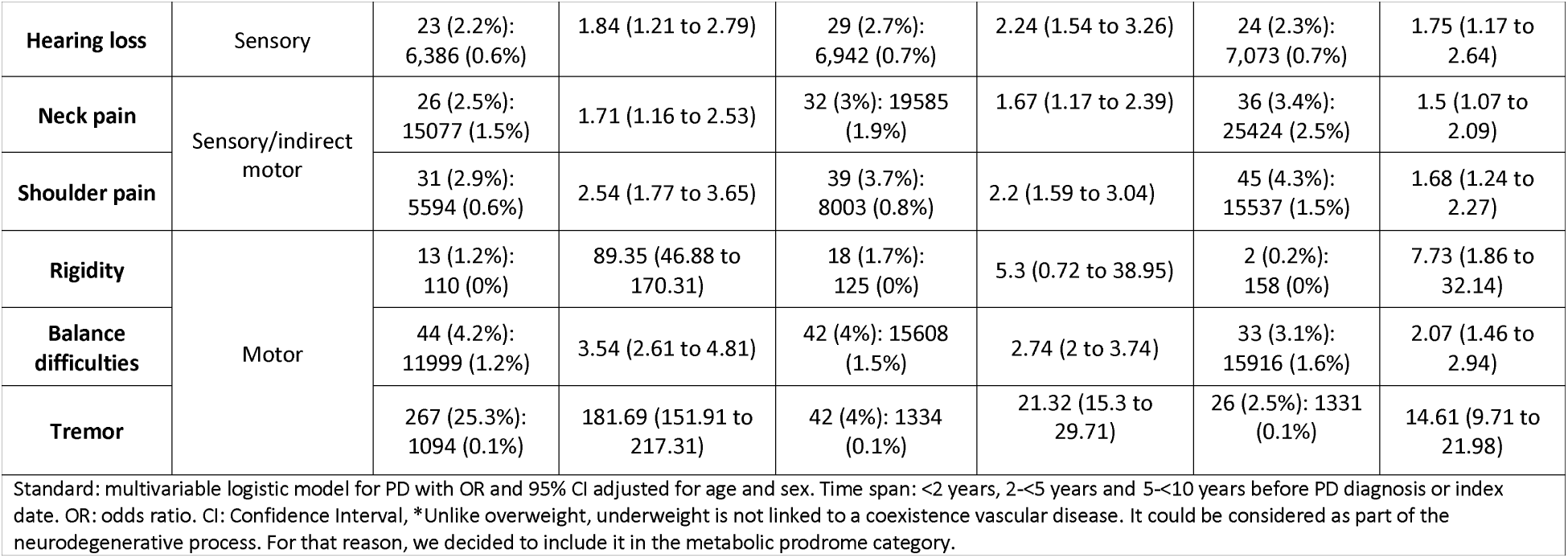
Prevalence and adjusted odds ratios of PD for prodromes and motor markers according to time of presentation.

#### Non-motor manifestations

Cognitive symptoms were the most frequently reported non-motor manifestations. Almost 5% of patients who were subsequently diagnosed with PD presented with ‘memory symptoms’, compared with <1% controls. Within 2 years of diagnosis, people with ‘memory symptoms’ had approximately 10-fold increased odds of PD (OR, 9.84; 95% CI, 7.39 to 13.11). This association remained up to 10 years prior to diagnosis (2-5 years: OR, 3.41; 95% CI, 2.13 to 5.46; 5-10 years: OR, 2.78; 95% CI, 1.38 to 5.61). People with hypotension had 7-fold increased odds of receiving PD diagnosis within 2 years (OR, 7.36; 95% CI, 4.2 to 12.93). This association remained at 2-5 years before diagnosis (OR, 6.22; 95% CI, 3.47 to 11.17).

Constipation was found to be more frequent amongst people with future PD compared to controls (4% vs 1%). After adjusting for age and sex, patients presenting to primary care with constipation were >3 times more likely to be diagnosed with PD compared to controls. This association remained stable over all three-time intervals (0-2 years: OR, 3.85; 95% CI, 2.84 to 5.24, 2-5 years: OR, 3.58; 95% CI, 2.71 to 4.74, 5-10 years: OR, 3.27; 95% CI, 2.47 to 4.33). The association between depression and subsequent PD was strongest up to 2 years prior to diagnosis of PD (OR, 4.47; 95% CI, 2.96 to 6.75), and the association weakened with greater time intervals (2-5 years: OR, 2.05; 95% CI, 1.28 to 3.27; 5-10 years: OR, 1.83; 95% CI, 1.25 to 2.67). Anxiety, fatigue, and insomnia had moderate associations with PD but, unlike depression, evidence for these associations was only present within 2 years from diagnosis. Overall, patients in East London presenting to primary care with psychiatric symptoms including depression, anxiety, fatigue, and insomnia within 2 years prior of diagnosis had about 2-4-fold increased odds of receiving a PD diagnosis (depression: OR, 4.47; 95% CI, 2.96 to 6.75, anxiety: OR, 3.13; 95% CI, 2.19 to 4.48; fatigue: OR, 2.14; 95% CI, 1.44 to 3.17; insomnia: OR, 2.29; 95% CI, 1.48 to 3.54) compared to those without.

Associations between pain and subsequent PD varied according to the location of pain. Shoulder pain was associated with a doubling of the odds of PD diagnosis up to 5 years prior to diagnosis (0-2 years: OR, 2.54; 95% CI, 1.77 to 3.65; 2-5 years: OR, 2.2; 95% CI, 1.59 to 3.04), whereas neck pain showed a weaker association with PD across the three periods (ORs ranging between 1.50 and 1.71). In contrast with other autonomic symptoms, such as constipation and hypotension, an association between erectile dysfunction and future PD was only observed 5-10 years prior to diagnosis (OR, 1.90; 95% CI, 1.46 to 2.61).

Hearing loss was more common in those who went on to develop PD (0-2 years: 2.2% vs 0.6%; 2-5 years: 2.7% vs 0.7%, 5-10 years: 2.3% vs 0.7%). Although the association with PD was weak, it was observed consistently across the three time periods (0-2 years: OR, 1.84; 95% CI, 1.21 to 2.79; 2-5 years: OR, 2.24; 95% CI, 1.54 to 3.26; 5-10 years: OR, 1.75; 95% CI, 1.17 to 2.64). Anosmia was present in less than 1% of PD cases across all time periods and was excluded from the analysis.

#### Motor manifestations

Tremor was the feature most strongly associated with subsequent PD across the three time periods. Most presentations of tremor were within 2 years of PD diagnosis with 25% individuals later diagnosed with PD having a record of tremor documented in their record compared with less than 1% of controls (OR, 181.69; 95% CI, 151.91 to 217.31). The prevalence of tremor was also higher in the PD group compared with controls in the second interval (2-5 years: OR, 21.32; 95% CI, 15.3 to 29.71) and third interval (5-10 years: OR, 14.61; 95% CI, 9.71 to 21.98) prior to diagnosis. Regarding other motor features, individuals who went on to receive PD diagnosis had a higher prevalence of balance difficulties across the three periods (4%) compared with controls (1%). Although rigidity was more frequently reported in the PD group than controls, it was not as common as other motor features with a prevalence of 1.2% during the first period (0-2 years) and <1% within the other two periods.

### Sensitivity analysis

We conducted a sensitivity analysis in which cases and controls were matched (1:10) on age and sex. There were no major differences between the matched and unmatched analysis, except for effect estimates for rigidity and balance difficulties. In terms of rigidity, there was a 45% relative change in ORs (matched-analysis OR, 130.0; 95% CI, 17.0 to 993.6, adjusted unmatched analysis OR, 89.3; 95% CI, 46.9 to 170.3) and 31.6% for balance difficulties (matched-analysis OR, 2.4; 95% CI, 1.7 to 3.4, adjusted unmatched analysis OR, 3.5; 95% CI 2.6 to 4.8) (TS1 in the supplementary material). However, the overall conclusions drawn from the sensitivity analysis were unchanged from the main analysis.

## Discussion

We used a large primary care dataset to explore risk factors and early clinical manifestations of PD in a highly diverse and generally deprived population. Based on the 2011 UK Census, London had the greatest ethnic diversity of anywhere in the UK, with the highest proportion of Black, South Asian and mixed/other ethnic groups, comprising ∼45% of local residents in East London compared with 14% in the rest of the UK. The fact that our study was undertaken in such a diverse setting, makes it easier to generalise our findings to the wider PD community.

There was no association between ethnic group or IMD and odds of PD. Although our findings need to be confirmed in other settings, the fact that no differences were found between groups suggests ethnicity and deprivation do not play a major role in PD risk, in contrast to what has recently been reported for dementia *[18]*. The fact that our findings are similar to studies undertaken in less diverse populations is reassuring and suggests that observations in other contexts have not introduced major selection bias.

We demonstrated that patients with PD manifest a range of motor and non-motor symptoms prior to diagnosis which might represent the prodromal stages of PD. Similar findings have been reported in more homogenous populations *[11,12,19]*, suggesting that non-specific clinical manifestations of PD may appear long before diagnosis can be established with high probability. In East London, we found that motor and cognitive symptoms were strongly associated with PD, up to 10 years before the diagnosis. Unlike other populations *[11,12]*, patients reporting memory complaints had a higher odds of being diagnosed with PD. The association remained even with presentations up to 10 years before diagnosis.

This is the first study focusing on the pre-diagnostic phase of PD in such a diverse population with universal access to healthcare. There are studies with a similar design but that have focused on less diverse populations. Plouvier and colleagues, in the Netherlands for example, carried out a nested case-control study using primary care data in a small sample size (86 cases and 78 controls) and a restricted period of 2 years before diagnosis *[10]*. They found that PD patients presented more often with functional complaints, autonomic symptoms (constipation and hyperhidrosis), and sleep problems than controls. Another study conducted in the Netherlands used primary care data to compare a group of 60 PD subjects and 58 controls *[11]*. They identified a pre-diagnostic period of 4-6 years which comprised a wide range of non-motor manifestations divided into 3 groups: psychological (mood disorders, dizziness), musculoskeletal (fibromyalgia, shoulder pain, radicular pain, low back pain, and arthralgia), and cardiorespiratory problems (hypertension, ischemic heart disease). Neither of these studies reported any motor manifestations before diagnosis. The THIN study shared a similar approach to the present study *[12]*. Both compared the medical records of a large sample of people with PD to healthy controls in a primary care setting using the same time frames (<2 years, 2-5 years, and 5-10 years) and had similar age and sex distribution. They differed by wealth and ethnicity, with the THIN database consisting mainly of a White and affluent participants compared to those residing in East London. The THIN data used prescribing data in addition to define the diagnostic groups including anxiety, depression, constipation, insomnia, and erectile dysfunction. Autonomic symptoms (constipation, hypotension, and erectile dysfunction) and mood disturbances (depression, anxiety, and insomnia) were on average between 2-4 times more commonly reported in THIN database than in East London across all periods studied (Table S3 supplementary material). Motor features, particularly tremor, were found to be the strongest markers of subsequent PD in both studies. Although tremor remained strongly associated to PD for a longer period prior to diagnosis (up to 10 years) in the THIN study. Prescribing data was not available in our study and may have contributed to the higher prevalence of certain symptoms in the THIN data. However, there may be other reasons for under-ascertainment of symptoms in East London. Minoritized ethnic and lower income groups may be more resilient or less likely to consult primary care physicians about symptoms such as constipation, fatigue, insomnia, and erectile dysfunction. A recent multicentric cross-sectional study of PD patients from a wide range of ethnicities (White, Black and Asian population) residing in London found that Asian PD patients scored higher in “sleep/fatigue” and “mood/apathy” questions on direct questioning *[20]*. Taken together these findings suggest non-motor symptoms might be under-reported amongst the East London population.

Epilepsy as a pre-diagnostic comorbidity of PD was observed in this setting, replicating observations from two other recent studies *[13,14]*. A role for PD as a risk factor for epilepsy has also been suggested *[21]*. Two observational study found that the prevalence of epilepsy in PD patients was higher than the estimated prevalence in the general population *[22,23]*, and in another study PD was associated with an increased risk of incident epilepsy, after adjusting for brain disorders, dementia, and antiepileptic drugs *[21]*. In our cohort, drug-induced parkinsonism could not be ruled out due to the lack of information about medication. Certain antiepileptic drugs have been associated with tremor and PD, such as valproate *[24]*, and co-existing vascular disease might link epilepsy with PD, especially in the elderly. Further research is needed to investigate a potential link.

In the East London population, having hypertension, T2D, and being underweight were associated with increased odds of developing PD. Our observations are in agreement with those of Kizza and colleagues *[25]*. Sauerbier and colleagues found that both Black and Asian patients with PD had higher prevalence of arterial hypertension and T2D, with brain imaging showing more pronounced white matter changes compared with White patients *[20]*. Of the vascular risk factors associated with an increase in PD risk, T2D is the one with the strongest evidence and anti-diabetic treatments such as metformin and exenatide are being investigated as potential treatments for PD *[26,27]*. Its role in PD risk and disease progression was explored in a recent meta-analyses of observational studies and genetic data *[28]*.

Hypertension has been associated with a small increase in the risk of PD according to a recent large meta-analysis, but a protective association was observed in a previous meta-analysis involving case-control studies only *[29]*. In the present study, being underweight was associated with an increased odds of future PD. There is conflicting evidence for BMI as a determinant of PD risk. A meta-analysis of prospective studies indicated no association *[30]*, but case-control studies consistently show an inverse association between BMI and PD *[31]*. A large prospective study in Korea found an inverse relationship between BMI and risk of PD, such that higher BMI was protective and lower BMI was a risk factor *[32]*. An additional challenge arises because weight loss is commonly encountered in the clinical course of PD and therefore some of the association may be driven by reverse causation. To try to mitigate this possibility members of our group showed a potential causal effect of lower BMI and increased risk of PD using Mendelian randomisation *[33]*.

Of the pre-diagnostic clinical manifestations, tremor showed the strongest association with subsequent PD, which was maintained even 10 years prior to diagnosis. Various studies support the idea that tremor may be an early feature of PD *[12]*. A longitudinal study conducted in Spain showed that after 3-years of follow-up, people with essential tremor were four times more likely to develop PD than those without tremor *[34]*. Similar results were found in another longitudinal study, where isolated tremor was associated with a doubling of the risk of PD *[35]*. Rigidity is a sign not a symptom explaining why it is rarely reported by patients. Indirect symptoms of rigidity (shoulder pain and neck pain) were more common in people with subsequent PD than controls, with an association were observed up to 10 years prior to diagnosis.

People with ‘memory problems’ had a higher odds of developing PD 10 years later. Compared to the THIN database, the association between cognitive symptoms and subsequent PD was stronger in the East London population, and could be a population-specific effect *[12]*. Black patients with PD have been reported as being more likely to have cognitive impairment *[1,20]*. In the aforementioned study, comparing Black and Asian patients to White patients with PD, it was found that Black people with PD had a greater cognitive impairment assessed by MMSE than White patients *[20]*. However, the method of assessment used may have produced artificially lower scores in certain groups due to language barriers and cultural interpretation of symptoms and psychological tests. It is notable that both ethnicity and deprivation appear to be determinants of dementia risk, which may explain the greater association between cognitive symptoms and PD in this setting *[18]*.

A greater proportion of subjects with a subsequent diagnosis of PD than controls had hearing loss up to 10 years before their diagnosis. Research has been carried out in relation to auditory processing difficulties in PD *[36,37]* and impaired recognition of musical and nonverbal vocal emotions *[38,39]*. Taking this finding together with the high prevalence of cognitive problems among the PD group, it is possible that hearing loss was associated with cognitive impairment. Depression and anxiety were more equally distributed among PD cases and controls in contrast to what has been reported in other cohorts *[10–12]*.

There are several limitations to this study. The main limitation is that these data are derived from routinely collected primary care data with under-ascertainment of factors of interest and high missingness. Although the year of recording for each variable was available, data are extracted in a cross-sectional manner, meaning that incidence rates could not be calculated. Another caveat to our study is that, although the National Health Service provides free care at the point of access to all patients, there may still be under ascertainment of PD cases. For example, there is preliminary evidence for atypical presentations in ethnic minority *[40]* and a higher likelihood of being (mis)labelled with vascular mimics of neurodegenerative disease *[1]*. Another limitation is the lack of information regarding medication which meant that it was not possible to create a more robust definition of PD, or include additional cases not recorded as PD but prescribed anti-parkinsonian medication or exclude cases with drug-induced parkinsonism. Ascertainment of exposure variables and risk factors may also be low, with mild or transient symptoms not being reported or recorded. Some symptoms lack context. For example, ‘memory problems’ recorded in primary care settings often lack supportive neurological examination or neuropsychological testing to support a formal diagnosis of cognitive impairment.

## Conclusions

This study shows that associations between comorbidities and early recognised pre-diagnostic symptoms of PD are demonstrable in a diverse, urban-dwelling population, with universal access to healthcare. Tremor and ‘memory symptoms’ occur up to 10 years before diagnosis had the strongest association. Further research is needed to explore the novel links between epilepsy and PD, as well as hearing loss and PD.

## Data Availability

The authors confirm that the data supporting the findings of this study are available within the article and its supplementary materials.

## Acknowledgement

We are grateful to the general practitioners and patients in East London for the use of data from their electronic health record and the Clinical Effectiveness Group, Queen Mary University of London who provided a de-identified curated extract of the relevant coded data. The code sets used were supported by the Secure Health Analysis and Research in East London (SHARE) study funded by Barts Charity and they are available from the authors.

## Ethnical and governance approvals

The general practitioners as data controllers approved the research uses of this data. The data used was based on routinely collected data in general practitioner electronic health records which is deidentified and published using aggregate counts and did not require ethics committee approval. The Clinical Effectiveness Group is the data processor, and the General Practices in the four CCGs are the data controllers. CEG has the written consent of all practices in the study area to use pseudonymised patient data for audit and research for patient benefit. The researchers adhere to the data protection principles of the Data Protection Act 2018, and all data was managed according to UK NHS information governance requirements. All outputs were in the form of aggregate patient data. The NHS Health Research Authority toolkit (http://www.hra-decisiontools.org.uk/ethics/) identified that Research Ethics Approval was not required for this project as all data is pseudonymised and presented in aggregate form. This was confirmed by the Chair of the North East London Strategic Information Governance Network.

All necessary patient/participant consent has been obtained and the appropriate institutional forms have been archived.

## References

1 Ben-Joseph A, Marshall CR, Lees AJ, et al. Ethnic Variation in the Manifestation of Parkinson’s Disease: A Narrative Review. J Parkinsons Dis 2020;10:31–45.

2 Siddiqui IJ, Pervaiz N, Abbasi AA. The Parkinson Disease gene SNCA: Evolutionary and structural insights with pathological implication. Sci Rep 2016;6:1–11.

3 Hall A, Bandres-Ciga S, Diez-Fairen M, et al. Genetic risk profiling in parkinson’s disease and utilizing genetics to gain insight into disease-related biological pathways. Int J Mol Sci 2020;21:1–15.

4 Nalls MA, Blauwendraat C, Vallerga CL, et al. Identification of novel risk loci, causal insights, and heritable risk for Parkinson’s disease: a meta-analysis of genome-wide association studies. Lancet Neurol 2019;18:1091–102.

5 Hemming JP, Gruber-Baldini AL, Anderson KE, et al. Racial and socioeconomic disparities in parkinsonism. Arch Neurol 2011;68:498–503.

6 Ritz B, Lee PC, Hansen J, et al. Traffic-related air pollution and parkinson’s disease in Denmark: A case–control study. Environ Health Perspect 2016;124:351–6.

7 Van Den Eeden SK, Tanner CM, Bernstein AL, et al. Incidence of Parkinson’s disease: Variation by age, gender, and race/ethnicity. Am J Epidemiol 2003;157:1015–22.

8 Abbas MM, Xu Z, Tan LCS. Epidemiology of Parkinson’s Disease—East Versus West. Mov Disord Clin Pract 2018;5:14–28.

9 GBD 2016 Parkinson’s Disease Collaborators. Global, regional, and national burden of Parkinson’s disease, 1990–2016: a systematic analysis for the Global Burden of Disease Study 2016. Lancet Neurol 2018;17:939–53.

10 Plouvier AOA, Hameleers RJMG, Van Den Heuvel EAJ, et al. Prodromal symptoms and early detection of Parkinson’s disease in general practice: A nested case-control study. Fam Pract 2014;31:373–8.

11 Gonera EG, Van’t Hof M, Berger HJC, et al. Symptoms and duration of the prodromal phase in Parkinson’s disease. Mov Disord 1997;12:871–6.

12 Schrag A, Horsfall L, Walters K, et al. Prediagnostic presentations of Parkinson’s disease in primary care: A case-control study. Lancet Neurol 2015;14:57–64.

13 Heilbron K, Noyce AJ, Fontanillas P, et al. The Parkinson’s phenome-traits associated with Parkinson’s disease in a broadly phenotyped cohort. npj Park Dis 2019;5.

14 Jacobs BM, Belete D, Bestwick J, et al. Parkinsons disease determinants, prediction and gene-environment interactions in the UK Biobank. 2020;:1046–54.

15 Noyce AJ, Bestwick JP, Silveira-Moriyama L, et al. Meta-analysis of early nonmotor features and risk factors for Parkinson disease. Ann Neurol 2012;72:893–901.

16 Schrag A, Anastasiou Z, Ambler G, et al. Predicting diagnosis of Parkinson’s disease: A risk algorithm based on primary care presentations. Mov Disord 2019;34:480–6.

17 Shetty K, Krishnan S, Thulaseedharan JV, et al. Asymptomatic Hearing Impairment Frequently Occurs in Early-Onset Parkinson’s Disease. J Mov Disord 2019;12:84–90.

18 Bothongo PL, Jitlal M, Parry E, et al. Ethnic and socioeconomic determinants of dementia risk: A nested case-control study in the population of East London. Alzheimer’s Dement 2020;16:37869.

19 Gaenslen A, Swid I, Liepelt-Scarfone I, et al. The patients’ perception of prodromal symptoms before the initial diagnosis of Parkinson’s disease. Mov Disord 2011;26:653–8.

20 Sauerbier A, Schrag A, Brown R, et al. Clinical non-motor phenotyping of black and Asian minority ethnic compared to white individuals with Parkinson’s disease living in the United Kingdom. J Parkinsons Dis 2021;11:299–307.

21 Gruntz K, Bloechliger M, Becker C, et al. Parkinson disease and the risk of epileptic seizures. Ann Neurol 2018;83:363–74.

22 Son AY, Biagioni MC, Kaminski D, et al. Parkinson’s Disease and Cryptogenic Epilepsy. Case Rep Neurol Med 2016;2016:1–4.

23 Gaitatzis A, Carroll K, Majeed A, et al. The epidemiology of the comorbidity of epilepsy in the general population. Epilepsia 2004;45:1613–22.

24 Zadikoff C, Munhoz RP, Asante AN, et al. Movement disorders in patients taking anticonvulsants. J Neurol Neurosurg Psychiatry 2007;78:147–51.

25 Kizza J, Lewington S, Mappin-Kasirer B, et al. Cardiovascular risk factors and Parkinson’s disease in 500,000 Chinese adults. Ann Clin Transl Neurol 2019;6:624–32.

26 Kim DS, Choi H Il, Wang Y, et al. A New Treatment Strategy for Parkinson’s Disease through the Gut–Brain Axis: The Glucagon-Like Peptide-1 Receptor Pathway. Cell Transplant 2017;26:1560–71.

27 Athauda D, Maclagan K, Skene SS, et al. Exenatide once weekly versus placebo in Parkinson’s disease: a randomised, double-blind, placebo-controlled trial. Lancet 2017;390:1664–75.

28 Chohan H, Senkevich K, Patel RK, et al. Type 2 Diabetes as a Determinant of Parkinson’s Disease Risk and Progression. Mov Disord 2021;36:1420–9.

29 Chen J, Zhang C, Wu Y, et al. Association between Hypertension and the Risk of Parkinson’s Disease: A Meta-Analysis of Analytical Studies. Neuroepidemiology 2019;52:181–92.

30 Wang YL, Wang YT, Li JF, et al. Body mass index and risk of Parkinson’s disease: A dose-response meta-analysis of prospective studies. PLoS One 2015;10.

31 van der Marck MA, Dicke HC, Uc EY, et al. Body mass index in Parkinson’s disease: a meta-analysis. Parkinsonism Relat Disord 2012;18:263–7.

32 Jeong SM, Han K, Kim D, et al. Body mass index, diabetes, and the risk of Parkinson’s disease. Mov Disord 2020;35:236–44.

33 Noyce AJ, Kia DA, Hemani G, et al. Estimating the causal influence of body mass index on risk of Parkinson disease: A Mendelian randomisation study. PLoS Med 2017;14:e1002314.

34 Benito-León J, Louis ED, Bermejo-Pareja F. Risk of incident Parkinson’s disease and parkinsonism in essential tremor: A population based study. J Neurol Neurosurg Psychiatry 2009;80:423–5.

35 Alarcón F, Maldonado J-C, Cañizares M, et al. Motor Dysfunction as a Prodrome of Parkinson’s Disease. J Parkinsons Dis 2020;10:1067–73.

36 Pisani V, Sisto R, Moleti A, et al. An investigation of hearing impairment in de-novo Parkinson’s disease patients: A preliminary study. Park Relat Disord 2015;21:987–91.

37 Folmer RL, Vachhani JJ, Theodoroff SM, et al. Auditory Processing Abilities of Parkinson’s Disease Patients. Biomed Res Int 2017;2017.

38 Lima CF, Garrett C, Castro SL. Not all sounds sound the same: Parkinson’s disease affects differently emotion processing in music and in speech prosody. J Clin Exp Neuropsychol 2013;35:373–92.

39 Hardy CJD, Marshall CR, Golden HL, et al. Hearing and dementia. J Neurol 2016;263:2339–54.

40 Chaudhuri KR, Hu MTM, Brooks DJ. Atypical parkinsonism in Afro-Caribbean and Indian origin immigrants to the UK. Mov Disord 2000;15:18–23.

