## Supplemental data for "Risk factors, early presentations, and clinical markers of Parkinson’s disease in Primary Care in a diverse UK population"

**Supplementary material**

| **Table 1. Sensitivity analysis matching 10 controls for each case according to age and sex** | | | | | | | |
| --- | --- | --- | --- | --- | --- | --- | --- |
|  | | **Time period** | | | | | |
|  |  | **<2 years** | | **2-<5 years** | | **5-<10 years** | |
| **Exposures** | **Category** | **Unadjusted**  **OR** | **Adjusted**  **OR** | **Unadjusted**  **OR** | **Adjusted**  **OR** | **Unadjusted OR** | **Adjusted**  **OR** |
| **Underweight*** | Metabolic | 2.73  (1.17 to 6.37) | 2.58  (1.1 to 6.02) | 0.81  (0.29 to 2.25) | 0.78  (0.28 to 2.18) | 1.45  (0.74 to 2.83) | (0.74 to 2.82) |
| **Hypotension** | Autonomic | 6.84  (3.38 to 13.9) | 6.81  (3.35 to 13.8) | 4.88  (2.44 to 9.77) | 4.73  (2.36 to 9.5) | 2.01  (0.83 to 4.85) | 1.9  (0.79 to 4.6) |
| **Constipation** |  | 3.29  (2.32 to 4.66) | 3.29  (2.32 to 4.67) | 2.68  (1.97 to 3.66) | 2.66  (1.95 to 3.63) | 2.96  (2.16 to 4.06) | 2.97  (2.16 to 4.07) |
| **Erectile dysfunction** |  | 1.14  (0.72 to 1.80) | 1.14  (0.72 to 1.8) | 1.13  (0.79 to 1.64) | 1.13  (0.78 to 1.63) | 1.51  (1.11 to 2.05) | 1.52  (1.12 to 2.08) |
| **Depression** | Neuropsychiatric | 4.69  (2.88 to 7.63) | 4.61  (2.82 to 7.52) | 1.65  (0.99 to 2.73) | 1.64  (0.98 to 2.72) | 1.97  (1.31 to 2.97) | 1.94  (1.29 to 2.92) |
| **Anxiety** |  | 3.08  (2.06 to 4.60) | 3.01  (2.02 to 4.5) | 1.32  (0.8 to 2.18) | 1.29  (0.78 to 2.13) | 1.55  (1.09 to 2.21) | 1.53  (1.07 to 2.18) |
| **Insomnia** |  | 2.18  (1.36 to 3.51) | 2.17  (1.35 to 3.48) | 1.85  (1.20 to 2.85) | 1.87  (1.21 to 2.88) | 1.48  (0.96 to 2.28) | 1.47  (0.95 to 2.27) |
| **Fatigue** |  | 1.91  (1.25 to 2.93) | 1.86  (1.21 to 2.85) | 1.41  (0.93 to 2.13) | 1.4  (0.93 to 2.11) | 1.08  (0.71 to 1.63) | 1.06  (0.7 to 1.6) |
| **Memory** |  | 8.6  (5.91 to 12.49) | 8.73  (6.0 to 12.7) | 3.08  (1.81 to 5.24) | 3.09  (1.81 to 5.26) | 2.06  (0.96 to 4.42) | 2.01  (0.93 to 4.31) |
| **Shoulder pain** | Sensory/indirect motor | 2.23  (1.5 to 3.3) | 2.25  (1.52 to 3.33) | 1.88  (1.33 to 2.66) | 1.89  (1.33 to 2.68) | 1.32  (0.96 to 1.81) | 1.31  (0.95 to 1.80) |
| **Neck pain** |  | 1.39  (0.92 to 2.11) | 1.39  (0.92 to 2.11) | 1.38  (0.95 to 2.01) | 1.37  (0.94 to 1.99) | 1.21  (0.85 to 1.72) | 1.21  (0.85 to 1.72) |
| **Tremor** | Motor | 146.0  (90.6 to 235.3) | 151.2  (93.7 to 244.0) | 14.5  (9.02 to 23.2) | 14.5  (9.02 to 23.3) | 11.7  (6.59 to 20.6) | 11.4  (6.43 to 20.2) |
| **Rigidity** |  | 130.0  (17.0 to 993.6) | 124.8  (16.3 to 956.4) | 2.5  (0.28 to 22.4) | 2.48  (0.28 to 22.3) | 10.0  (1.41 to 71.0) | 8.27 (1.14 to 59.8) |
| **Balance difficulties** |  | 2.42  (1.73 to 3.39) | 2.4  (1.71 to 3.36) | 2.14  (1.52 to 3.01) | 2.1  (1.49 to 2.95) | 1.51  (1.04 to 2.19) | 1.49  (1.02 to 2.17) |
| *OR adjusted for ethnicity and IMD. | | | | | | | |

| **Table 2. Prevalence* of motor symptoms (tremor, rigidity or balance difficulties) and cognitive symptoms amongst PD patients according to ethnicity together with their association with future PD** | | | | | | |
| --- | --- | --- | --- | --- | --- | --- |
|  | **Motor symptoms** | | | **Cognitive symptoms** | | |
| **Ethnicity** | **Absent** | **Present** | **OR (95% CI)** | **Absent** | **Present** | **OR (95% CI)** |
| **White** | 323 (60%) | 214 (40%) | 1 | 493 (92%) | 43 (8%) | 1 |
| **Black** | 93 (56%) | 73 (44%) | 1.18 (0.83 to 1.68) | 153 (92%) | 13 (8%) | 0.97 (0.51 to 1.86) |
| **S Asian** | 108 (52%) | 100 (48%) | 1.40 (1.01 to 1.93) | 186 (89%) | 22 (11%) | 1.36 (0.79 to 2.33) |
| **Other** | 51 (58%) | 37 (42%) | 1.10 (0.69 to 1.73) | 73 (84%) | 14 (16%) | 2.20 (1.15 to 4.22) |
| **Unknown** | 43 (77%) | 13 (23%) | 0.46 (0.24 to 0.87) | 54 (96%) | 2 (4%) | 0.42 (0.10 to 1.80) |
| *The proportion covered the whole 10 year period studied | | | | | | |

| **Table 3. Distribution of pre-diagnostic manifestation amongst PD and controls in the ELGP and THIN database** | | | | | | | |
| --- | --- | --- | --- | --- | --- | --- | --- |
|  | | **Time period** | | | | | |
| **Exposures** | **Category** | **<2 years** | | **2-<5 years** | | **5-<10 years** | |
|  |  | East London  (%) | THIN  (%) | East London  (%) | THIN  (%) | East London  (%) | THIN  (%) |
| **Hypotension** | Autonomic | 1.2 | 2.1 | 1.1 | 1.6 | 0.6 | 1.6 |
| **Constipation** |  | 4.2 | 32.2 | 5 | 25.1 | 5 | 19.9 |
| **Erectile dysfunction** |  | 2.0 | 7.0 | 3.2 | 8.0 | 4.8 | 11.0 |
| **Depression** | Neuropsychiatric | 2.3 | 9.6 | 1.7 | 6.5 | 2.7 | 5.6 |
| **Anxiety** |  | 3.0 | 8.6 | 1.7 | 7.0 | 3.5 | 8.1 |
| **Insomnia** |  | 2.0 | 4.3 | 2.4 | 3.9 | 2.3 | 5.1 |
| **Fatigue** |  | 2.5 | 10.5 | 2.6 | 9.0 | 2.5 | 10.7 |
| **Memory** |  | 4.9 | 2.7 | 1.7 | 1.3 | 0.8 | <1 |
| **Shoulder pain** | Sensory/indirect motor | 2.9 | 7.3 | 3.7 | 8.5 | 4.3 | 10.4 |
| **Neck pain** |  | 2.5 | 4.2 | 3.0 | 4.7 | 3.4 | 7.6 |
| **Tremor** | Motor | 25.3 | 40.7 | 4.0 | 6.5 | 2.5 | 1.7 |
| **Rigidity** |  | 1.2 | 2.8 | <1 | <1 | <1 | <1 |
| **Balance difficulties** |  | 4.2 | 4.1 | 4.0 | 1.6 | 3.1 | 1.2 |
